# Opioid Use Disorder is Associated with Lower Resting-State Brain Network Segregation

**DOI:** 10.64898/2026.08.04.26359717

**Authors:** Delaney McKinstry, Xinyi Li, Astrid P. Ramos-Rolón, Nathan M. Hager, Samnang T. Kim, Nora A. Foster, Timothy Pond, Lindsey M. Brier, Daniel D. Langleben, Anna Rose Childress, Henry R. Kranzler, Jacob G. Dubroff, Ilya M. Nasrallah, W. Andrew Kofke, Paul Regier, Corinde E. Wiers, Zhenhao Shi

**Author notes:** Correspondence: Corinde E. Wiers, PhD, 3535 Market St Ste 500, Philadelphia, PA 19104, Zhenhao Shi, PhD, 3535 Market St Ste 500, Philadelphia, PA 19104. The authors contributed equally (Corinde E. Wiers, Zhenhao Shi).

## Abstract

**Background:** Opioid use disorder (OUD) is associated with a wide range of cognitive, affective, and motivational impairments, suggesting a disruption of large-scale brain systems that support diverse domains of functioning. Resting-state brain network segregation quantifies the degree of functional specialization within brain networks, is age-related, has been linked to brain glucose metabolism, and has been shown to be reduced in substance use disorders. We examined brain network segregation in individuals with OUD and non-OUD controls and tested associations with the duration of opioid use.

**Methods:** Resting-state functional MRI data were collected from 149 individuals with OUD and 126 non-OUD controls. Functional connectivity was computed between brain regions assigned to functionally specialized networks supporting higher-order “association” or “sensorimotor” processes. For each network, segregation was quantified as the extent to which within-network connectivity exceeded between-network connectivity.

**Results:** Individuals with OUD demonstrated lower segregation of the association and sensorimotor networks than non-OUD controls. Within the OUD group, more years of opioid use was associated with lower segregation of the association network, but not the sensorimotor network.

**Conclusions:** OUD is characterized by overall lower resting-state brain network segregation. More years of opioid exposure was associated with lower association-network segregation, consistent with there being cumulative effects of chronic opioid use on large-scale brain organization, though causation could not be examined in this cross-sectional dataset. These findings identify altered network segregation as a potential neurobiological marker of OUD and suggest that restoration of brain network specialization is a measurable target of OUD treatment and potentially recovery.

## INTRODUCTION

Opioid use disorder (OUD) is a major public health crisis, affecting millions of individuals in the United States each year, with opioid-related overdoses due largely to illicit fentanyl. OUD is also associated with broad impairments in executive functioning, attention, working memory, emotion regulation, and decision-making, suggesting that a widespread disruption of higher-order neural systems is involved in cognitive control ^1–4^. Characterizing the large-scale neural organization associated with OUD could improve our understanding of the neurobiological mechanisms underlying the disorder and help to identify novel therapeutic targets.

Resting-state functional connectivity measured with functional magnetic resonance imaging (fMRI) has emerged as a powerful tool for examining large-scale brain network organization in substance use disorders (SUDs) ^5–11^. Resting-state networks represent distributed brain systems defined by spontaneous neural activity and are thought to reflect functional architecture underlying cognition and behavior. Prior work in OUD and other SUDs has consistently demonstrated dysfunction in higher-order the association network, particularly the default mode network, frontoparietal network, salience network, and attention-related systems ^11–15^. In addition to association systems, sensorimotor and visual networks may also be disrupted in OUD, potentially reflecting the widespread effects of chronic opioid exposure on sensory processing and motor function ^16–19^.

One approach for characterizing large-scale brain organization is network segregation, a measure of the extent to which functional networks maintain distinct patterns of connectivity. Brain network segregation is typically quantified by comparing the strength of within-network connectivity to connectivity between networks ^20^. Higher segregation indicates greater within-network connectivity and that networks are more functionally specialized, whereas lower segregation reflects greater inter- network integration and less network distinctiveness. A balance between segregation and integration is thought to support efficient information processing by allowing specialized neural systems to operate independently while remaining capable of coordinated communication when necessary ^21^.

Alterations in brain network segregation have been associated with aging, cognitive decline, and several neuropsychiatric conditions ^22–28^. Across the adult lifespan, resting-state networks become progressively less segregated, which has been linked to poorer cognitive and motor performance ^29–31^. There is emerging evidence that similar patterns of reduced network specialization may characterize SUDs. For example, individuals with alcohol use disorder exhibit lower segregation across association and sensorimotor networks ^32^. Consistent with this framework, our group recently reported that among individuals with OUD, greater drug use severity is associated with lower segregation of the fronto- parietal and salience networks ^16^. These findings suggest that chronic substance use may disrupt the boundaries between functionally specialized brain systems.

In the present study, we investigated resting-state brain network segregation in a pooled sample of individuals with OUD and in non-OUD control participants. Given prior evidence of altered brain network connectivity in OUD, we hypothesized that individuals with OUD would exhibit lower brain network segregation than controls, particularly within the higher-order association network. We also examined whether the duration of opioid use was associated with segregation within these systems, potentially reflecting cumulative effects of chronic opioid exposure on large-scale brain organization.

## METHODS

### Participants

A total of 149 patients with OUD and 126 non-OUD controls who completed structural MRI and resting- state fMRI were included in the study. Participants were drawn from multiple studies conducted at the Center for Studies of Addiction at the University of Pennsylvania ^33–37^. Across studies, common inclusion criteria for both groups were the ability to read and speak English, good physical health as determined by medical history and physical examination, and no contraindications to structural or functional MRI (e.g., claustrophobia, indwelling magnetically active foreign bodies, history of clinically significant head trauma, or use of medications that could confound blood oxygen level-dependent [BOLD] brain responses). For the OUD group, additional inclusion criteria were a DSM-IV-TR diagnosis of current opioid dependence, a DSM-5 diagnosis of current OUD, or evidence of current treatment with medication for OUD. For the non-OUD control group, exclusion criteria included a history of a DSM-IV- TR opioid dependence or DSM-5 OUD diagnosis. See the **Supplementary Materials** for study-specific eligibility criteria. All studies were approved by the University of Pennsylvania Institutional Review Board. Written informed consent was obtained from all participants, who were paid for their participation, and all procedures were conducted in accordance with relevant guidelines and regulations.

### MRI Data Acquisition and Processing

MRI was performed on Siemens (Siemens, Erlangen, Germany) TIM Trio and Prisma 3T systems. Blood oxygenation level-dependent resting-state fMRI images were acquired using a whole-brain, single-shot gradient-echo echo-planar sequence. High-resolution structural MRI images were acquired using a multi-echo magnetization-prepared rapid acquisition gradient-echo sequence. See the **Supplementary Materials** for study-specific MRI acquisition parameters.

The resting-state fMRI data were preprocessed in MATLAB using a pipeline adapted from Ciric and colleagues (2018) ^38^. This consisted of removing the first ten seconds of data, estimation of the 24 motion parameters (including the six raw motion parameters, six framewise displacement parameters, and the square of the raw motion and framewise displacement parameters), removal of individuals with mean framewise displacement>0.5 mm, slice time correction, motion correction, coregistration and segmentation of the structural images, skull stripping, computation of DVARS and identification of DVARS outliers using the procedure described in Afyouni and Nichols ^39^, despiking using AFNI’s 3dDespike, removal of polynomial trends, extraction of nuisance signals from the voxels located within the top 10% of the deepest tissue of the white matter and the cerebrospinal fluid, interpolation of framewise displacement>0.5mm and DVARS outlier time points using Lomb-Scargle periodogram, bandpass filtering at 0.01–0.1 Hz of the images and covariates (i.e., 24 motion parameters and two nuisance signals), regressing out the filtered covariates, spatial smoothing using a Gaussian kernel with a full width at half maximum of 8 mm, and spatial normalization to the Montreal Neurological Institute (MNI) space.

### Quantification of Brain Network Metrics

We followed the procedure of previous brain network segregation studies ^16,32,40^ and defined 264 spherical brain regions of interest (ROIs) using the Power-264 atlas ^41^. Based on the atlas, the ROIs were assigned to one of 13 distinct functional brain networks, which were further grouped into three categories: (1) “association” network (i.e., cingulo-opercular, dorsal attention, default mode, fronto- parietal, salience, and ventral attention); (2) “sensorimotor” network (i.e., auditory, hand, mouth, and visual); and (3) “other” networks (i.e., cerebellar, memory, and subcortical) ^16,32,40,41^.

For each participant, an adjacency matrix was generated by calculating the Fisher Z-transformed Pearson correlation coefficients between time courses of the ROIs. The segregation of each brain network was computed as (*̅Z*_w_ − *̅Z_b_*)/*̅Z*_w_, where *̅Z*_w_ is the mean Z-transformed correlation coefficient between the ROIs within that network (i.e., within-network connectivity), and *̅Z_b_* the mean coefficient between the ROIs of that network and the ROIs of other networks (i.e., between-network connectivity). Only positive coefficients were used in the calculations to avoid spurious contributions of negative correlations ^16,32,40,41^. At the network category level, we calculated segregation of the sensorimotor and association networks by averaging the network segregation of their constituent networks. We also computed a segregation score for each ROI using the same approach, based on its individual connectivity with other ROIs within the same network versus those of other networks.

We conducted exploratory graph-theoretical analyses to aid the interpretations of the observed group difference in brain network segregation. First, we examined whether such differences persisted without using an atlas to prespecify network assignments. To this end, we calculated the modularity index, a data-driven counterpart of network segregation, from each participant’s individualized network partition. Second, we examined whether reduced network segregation could be attributed to alternative brain network topology that deviated from the canonical Power-264 template. To quantify such deviation, we calculated the normalized mutual information to measure the similarity between the canonical Power- 264 template and an empirically derived, data-driven network partition for each participant. Lastly, we explored whether lower brain network segregation was accompanied by greater whole-brain integration by calculating global network efficiency. Detailed procedures for the calculation of modularity, normalized mutual information, and global efficiency are provided in the **Supplementary Materials**.

### Statistical Analysis

Statistical analysis was conducted in R (version 4.5.2). Two-sample t-tests and χ² tests were conducted to compare participant characteristics between the OUD and non-OUD groups for numerical and categorical variables, respectively. Linear mixed-effects models ^42,43^ were used to compare network segregation between the OUD and non-OUD groups while adjusting for sex, age, race, tobacco use, stimulant use, scanner parameters, mean framewise displacement, and study-specific random intercepts. Within the association network, p-values were corrected using a false discovery rate (FDR) across the analyses of six individual networks. The same correction was performed across the four individual sensorimotor networks. Detailed statistical results such as uncorrected p-values, effect sizes, and marginal effects are reported in the **Supplementary Materials**.

Exploratory analysis examined the association between years of opioid use and network segregation while adjusting for the aforementioned covariates. Other exploratory analysis examined the effects of sex, age, history of overdose, and type of medication for OUD on network segregation, which are reported in the **Supplementary Materials**.

## RESULTS

The demographic and clinical characteristics of the participants are summarized in **Table 1**. Variables that differed significantly between the OUD and non-OUD groups (i.e., age, race, tobacco use, stimulant use, and framewise displacement) were included as covariates in all subsequent analyses to account for potential confounding effects.

**Table 1.**
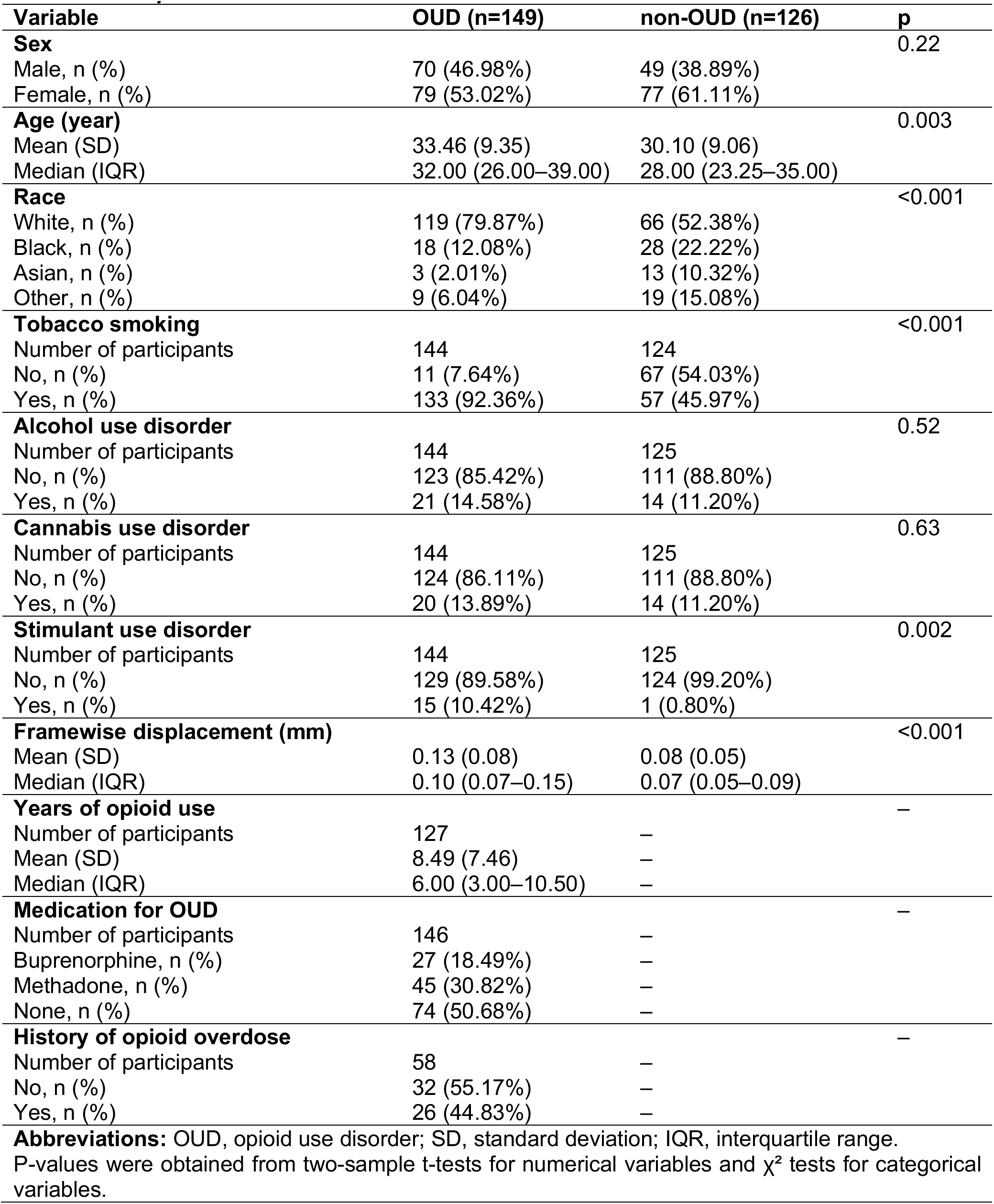
Participant characteristics.

| Variable | OUD (n=149) | non-OUD (n=126) | p |
| --- | --- | --- | --- |
| **Sex** |  |  | 0.22 |
| Male, n (%) | 70 (46.98%) | 49 (38.89%) |  |
| Female, n (%) | 79 (53.02%) | 77 (61.11%) |  |
| **Age (year)** |  |  | 0.003 |
| Mean (SD) | 33.46 (9.35) | 30.10 (9.06) |  |
| Median (IQR) | 32.00 (26.00–39.00) | 28.00 (23.25–35.00) |  |
| **Race** |  |  | <0.001 |
| White, n (%) | 119 (79.87%) | 66 (52.38%) |  |
| Black, n (%) | 18 (12.08%) | 28 (22.22%) |  |
| Asian, n (%) | 3 (2.01%) | 13 (10.32%) |  |
| Other, n (%) | 9 (6.04%) | 19 (15.08%) |  |
| **Tobacco smoking** |  |  | <0.001 |
| Number of participants | 144 | 124 |  |
| No, n (%) | 11 (7.64%) | 67 (54.03%) |  |
| Yes, n (%) | 133 (92.36%) | 57 (45.97%) |  |
| **Alcohol use disorder** |  |  | 0.52 |
| Number of participants | 144 | 125 |  |
| No, n (%) | 123 (85.42%) | 111 (88.80%) |  |
| Yes, n (%) | 21 (14.58%) | 14 (11.20%) |  |
| **Cannabis use disorder** |  |  | 0.63 |
| Number of participants | 144 | 125 |  |
| No, n (%) | 124 (86.11%) | 111 (88.80%) |  |
| Yes, n (%) | 20 (13.89%) | 14 (11.20%) |  |
| **Stimulant use disorder** |  |  | 0.002 |
| Number of participants | 144 | 125 |  |
| No, n (%) | 129 (89.58%) | 124 (99.20%) |  |
| Yes, n (%) | 15 (10.42%) | 1 (0.80%) |  |
| **Framewise displacement (mm)** |  |  | <0.001 |
| Mean (SD) | 0.13 (0.08) | 0.08 (0.05) |  |
| Median (IQR) | 0.10 (0.07–0.15) | 0.07 (0.05–0.09) |  |
| **Years of opioid use** |  |  | – |
| Number of participants | 127 | – |  |
| Mean (SD) | 8.49 (7.46) | – |  |
| Median (IQR) | 6.00 (3.00–10.50) | – |  |
| **Medication for OUD** |  |  | – |
| Number of participants | 146 | – |  |
| Buprenorphine, n (%) | 27 (18.49%) | – |  |
| Methadone, n (%) | 45 (30.82%) | – |  |
| None, n (%) | 74 (50.68%) | – |  |
| **History of opioid overdose** |  |  | – |
| Number of participants | 58 | – |  |
| No, n (%) | 32 (55.17%) | – |  |
| Yes, n (%) | 26 (44.83%) | – |  |

**Abbreviations:** OUD, opioid use disorder; SD, standard deviation; IQR, interquartile range.
P-values were obtained from two-sample t-tests for numerical variables and $\chi^2$ tests for categorical variables.

We found a significant effect of group on brain network segregation of the association network (F(1,241.57)=12.27, p<0.001), with OUD participants showing lower segregation (0.25, 95% confidence interval=[0.22,0.27]) than the non-OUD group (0.29 [0.26,0.31]) (see **Figure 1A**). Further examination of individual association networks showed lower segregation in the OUD group than the non-OUD group in the cingulo-opercular, default mode, and ventral attention networks (pFDR=0.003–0.026; see **Figure S1**). There were no group differences in brain network segregation of the dorsal attention, fronto- parietal, and salience networks (pFDR=0.13–0.92; see **Figure S1**). See details in **Supplementary Table S1**.

**Figure 1.**
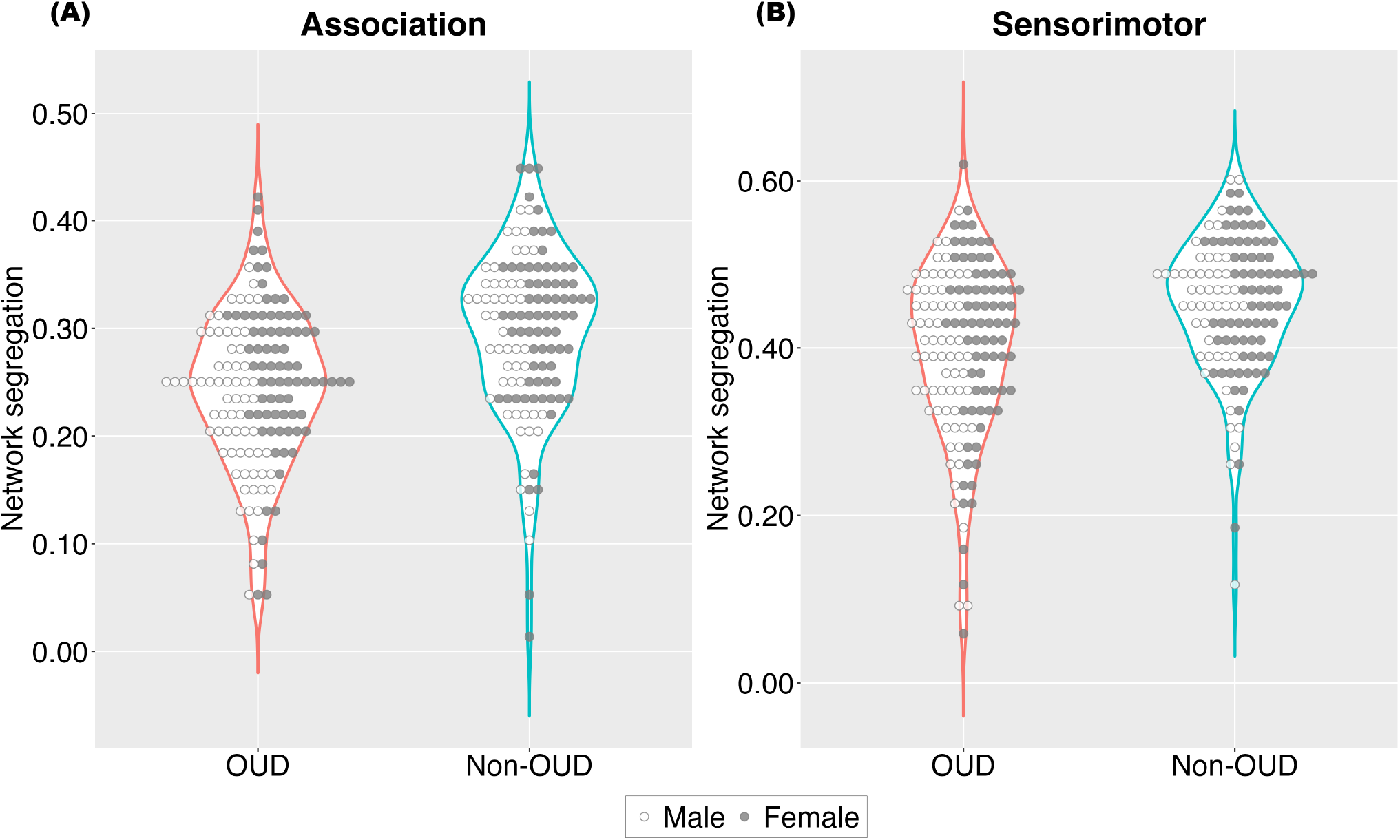
Lower brain network segregation in the OUD compared to the non-OUD individuals in the association and sensorimotor networks. Abbreviation: OUD, opioid use disorder.

We also found a significant effect of group on brain network segregation of the sensorimotor network (F(1,209.83)=13.14, p<0.001), such that the OUD group showed lower segregation (0.41 [0.37,0.45]) than the non-OUD group (0.46 [0.42,0.50]) (see **Figure 1B**). Further examination of individual sensorimotor networks showed lower segregation in the OUD group than the non-OUD group in the hand and visual networks (pFDR=0.005–0.031; see **Figure S1**). There were no group differences in network segregation of the auditory and mouth networks (pFDR=0.12; see **Figure S1**). See details in **Supplementary Table S1**.

We examined the segregation scores of individual ROIs within the networks that exhibited significant group difference in network segregation. Compared to the non-OUD group, the OUD group had lower segregation scores in the dorsal anterior cingulate cortex, the right anterior insular cortex, the bilateral precentral and postcentral gyri, the bilateral middle temporal gyrus, and the bilateral fusiform gyrus (pFDR<0.05).

Within the OUD group, there was a significant association between years of opioid use and segregation of the association network (F(1,115.00)=4.05, p=0.046), such that patients with longer opioid use history had lower brain network segregation (slope=–2.21 [–4.27,–0.15] ×10⁻³; see **Figure 2A**). Among the individual association networks, the negative association was found in the default mode and dorsal attention networks, although the statistical significance did not survive the FDR correction (p=0.029– 0.035, pFDR=0.10; see **Figure S2**). The association with years of opioid use was not significant for the sensorimotor network (F(1,114.25)=1.25, p=0.27; 1.58 [–0.66,4.72] ×10⁻³; see **Figure 2B**) or the individual networks within it (p=0.17–0.94; see **Figure S2**). See details in **Supplementary Table S2**.

**Figure 2.**
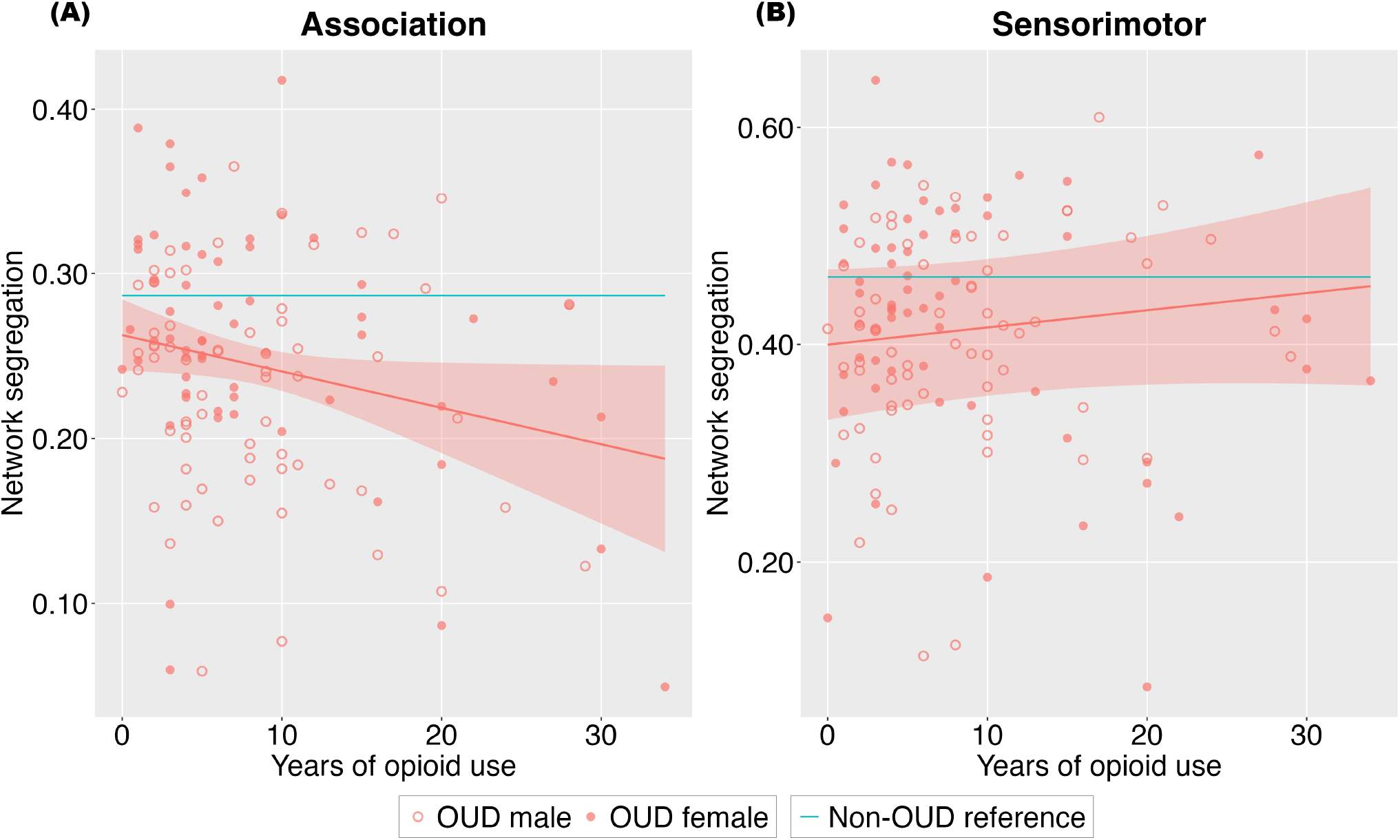
Negative association between years of opioid use and network segregation for the association network but not the sensorimotor network. Shaded areas represent 95% confidence intervals. Abbreviation: OUD, opioid use disorder.

Consistent with prior studies, there was a main effect of age, which was negatively associated with brain network segregation of both the association network (F(1,134.41)=20.19, p<0.001; slope=–2.14 [– 3.09,–1.28], ×10⁻³) and the sensorimotor network (F(1,201.85)=9.89, p=0.002; –1.90 [–2.62,–0.37], ×10⁻³) (see **Supplementary Table S3** and **Figure S3**). There was no significant interaction between group and age on segregation of either the association network or the sensorimotor network (p=0.28– 0.72) (see **Table S4**).

Exploratory graph-theoretical analyses showed a significant effect of group on brain network modularity (F(1,237.17)=4.86, p=0.029), such that the OUD group had lower modularity (0.26 [0.24,0.28]) than the non-OUD group (0.27 [0.26,0.29]). There was also a significant group effect on normalized mutual information (F(1,244.87)=14.78, p<0.001), such that the OUD group showed lower similarity between individualized brain partition and the Power-264 template (0.15 [0.14,0.16]) than the non-OUD group (0.17 [0.16,0.18]). The OUD and non-OUD groups did not differ on brain network integration as indexed by the global efficiency metric (F(1,245.54)=0.80, p=0.37; OUD, 0.62 [0.62,0.63]; non-OUD, 0.62 [0.62,0.63]).

## DISCUSSION

We compared resting-state brain network segregation in individuals with OUD and non-OUD controls. Individuals with OUD showed significantly lower brain network segregation of the association network than non-OUD controls, particularly within the default mode, cingulo-opercular, and ventral attention networks. Lower segregation was also observed within the sensorimotor network, specifically in the hand and visual networks. In addition, longer duration of opioid use was associated with lower segregation in the association network, particularly the default mode and dorsal attention networks, but not the sensorimotor network. Age was also negatively associated with segregation across many networks, consistent with previous studies demonstrating age-related reductions in brain network specialization ^44^. Exploratory analyses also showed lower brain modularity and greater deviation from the canonical network template in individuals with OUD. Furthermore, the lower brain network segregation in individuals with OUD did not translate into greater brain network integration. These findings extend our group’s prior work demonstrating associations between greater drug use severity and lower segregation in the attention network in individuals with OUD seeking naltrexone treatment ^16^. The present study expands those observations in a substantially larger and more heterogeneous cohort, and, importantly, includes a non-OUD comparison group. Together, these findings provide convergent evidence that OUD is associated with widespread alterations in large-scale brain organization.

The most robust reductions in brain network segregation were observed within the default mode, cingulo-opercular, and ventral attention networks. The default mode network supports internally directed cognition, including self-referential processing, autobiographical memory, and future-oriented thinking ^45–47^. Previous rsFC studies have consistently reported altered default mode network connectivity in OUD ^14,15,48^. In individuals with SUDs, the default mode network has been shown to facilitate drug craving and relapse ^49,50^. Thus, lower default mode network segregation may contribute to maladaptive self-referential processing, altered decision-making, and drug use. Similarly, the cingulo-opercular network plays a central role in sustained cognitive control, performance monitoring, and maintenance of task goals, while the ventral attention network is involved in detecting behaviorally salient stimuli and reorienting attention ^51–55^. Lower segregation within these networks may indicate less efficient cognitive control and attentional processes, consistent with the altered executive functioning that has been repeatedly observed in OUD and other SUDs ^56^. Finally, we observed an association between longer opioid use duration and lower segregation of the association network, suggesting that prolonged opioid exposure may lead to progressive brain network alterations and providing convergent validation of the main findings. Thus, OUD appears to be associated with widespread reductions of large-scale brain networks that support internally and externally directed cognition, cognitive control, and attentional orienting, potentially contributing to altered behavior and cognition in OUD.

In addition to the association network, individuals with OUD exhibited lower segregation within sensorimotor systems, particularly the hand and visual networks. Although research in SUD populations has traditionally focused on higher-order cognitive networks, accumulating evidence suggests that chronic opioid use may also affect more basic sensory and motor systems ^9,19,57^. These findings indicate that network-level alterations in OUD are not confined to executive control systems but may extend across multiple levels of neural organization, reflecting broad effects on brain function.

The present findings may also relate to brain energetics. Brain network segregation has been linked to cerebral glucose metabolism measured with ¹⁸F-fluorodeoxyglucose positron emission tomography ([¹⁸F]FDG PET), with lower segregation associated with lower glucose utilization and poorer cognitive performance in aging ^40^. Consistent with this framework, aging is characterized by reductions in both cerebral glucose metabolism and network segregation, and we similarly observed age-related reductions in segregation across multiple networks. Furthermore, individuals with OUD have been reported to exhibit widespread reductions in cerebral glucose metabolism relative to controls ^58–60^. Thus, the lower network segregation observed in OUD may reflect altered metabolic demand and diminished energetic support for maintaining functionally specialized neural systems. This interpretation is consistent with growing evidence implicating disrupted brain energetics in addiction and suggests that altered metabolism may contribute to large-scale functional network reorganization in OUD ^61^. Future multimodal studies combining resting-state fMRI and FDG PET will be needed to directly test this hypothesis.

There are limitations that should be considered when interpreting these findings. First, the cross-sectional design precludes causal inference. Although lower network segregation was observed in individuals with OUD and was associated with a longer duration of opioid use, the present findings cannot distinguish whether altered network organization is a consequence of chronic opioid exposure or a pre-existing neurobiological vulnerability that increases susceptibility to opioid use and the development of OUD. Longitudinal studies spanning opioid initiation, active use, treatment, and recovery will be necessary to disentangle predisposing factors from the effects of chronic opioid exposure. Second, the study data were pooled across seven separate studies, with heterogeneous recruitment procedures, scanner parameters, and clinical characteristics. Individuals with OUD also frequently use other substances and have co-occurring psychiatric, psychosocial, and other health-related factors that may influence functional connectivity ^62,63^. By correcting for many of the group differences, we increased confidence that the observed findings reflect features associated with OUD.

In conclusion, individuals with OUD demonstrated widespread differences from controls in brain network segregation across both the association and sensorimotor networks, which was not accompanied by an increase in network integration, and longer opioid use duration was associated with lower segregation of the association network. These findings suggest that OUD is associated with reduced functional specialization and altered large-scale brain organization, potentially reflecting altered brain energetics, diminished neural efficiency, and processes that overlap with age-related declines in network organization. Restoration of healthy brain network organization may represent a measurable target for future interventions in OUD.

## Data Availability

The data underlying this article can be shared on reasonable request to the corresponding authors.

## Acknowledgements

This work was supported by the following National Institutes of Health grants: DA051709 (Shi), DA046345 (Wiers, Dubroff, Nasrallah, Pond, Kranzler), DA051737 (Regier), DA036028 (Langleben), DA028874 (Li, Hager), AG076411 (Ramos-Rolón). It was also supported by the Commonwealth Fund of Pennsylvania CURE grant SAP#4100055577 (Langleben, Childress) and the NARSAD Young Investigator Grant from the Brain & Behavior Research Foundation (#30780, Shi).

## Data Availability

The data underlying this article can be shared on reasonable request to the corresponding authors.

