## Supplementary Materials for "Opioid Use Disorder is Associated with Lower Resting-State Brain Network Segregation"

*The authors contributed equally.

#### Table of Contents

SUPPLEMENTARY METHODS ........ ***s2***
- Study-specific eligibility criteria and MRI acquisition parameters........ s2
- Exploratory graph-theoretical analyses........ s5

SUPPLEMENTARY RESULTS........ ***s7***
- Effects of sex, overdose, and medication ........ s7

SUPPLEMENTARY TABLES........ ***s8***
- Table S1. Main effect of group........ s8
- Table S2. Main effect of years of opioid use........ s8
- Table S3. Main effect of age (years)........ s9
- Table S4. Interaction between group and age (years)........ s9
- Table S5. Main effect of sex........ s10
- Table S6. Interaction between group and sex. ........ s10
- Table S7. Main effect of opioid overdose history. ........ s11
- Table S8. Main effect of medication........ s11

SUPPLEMENTARY FIGURES ........ ***s12***
- Figure S1. Segregation of individual brain networks........ s12
- Figure S2. Association between years of opioid use and segregation of individual brain networks........ s13
- Figure S3. Association between age and segregation of the association and sensorimotor networks. ........ s14

REFERENCES........ ***s15***

### SUPPLEMENTARY METHODS

#### *Study-specific eligibility criteria and MRI acquisition parameters*

**Study 1** (11 OUD, 24 non-OUD). Eligibility criteria for both groups: 18–60 years of age; no claustrophobia or other MRI contraindications; working command of English; a negative HIV test at screening; not currently pregnant or breastfeeding; body girth ≤52 in, head girth ≤25 in, and body weight ≤500 lbs; no seizure disorder; no history of significant head trauma, stroke, or stroke-related spasticity; no history of schizophrenia, schizoaffective disorder, or bipolar disorder type 1; no illicit opioid use in the past week or positive urine drug test for illicit opioids; no medical or psychological condition that may compromise participant safety or successful participation in the study; no vision problems that cannot be corrected with glasses. Additional eligibility criteria for the OUD group: DSM-5 diagnosis of OUD; treatment with a stable dosage of medication for OUD; reporting opioids as the drug of choice. Additional eligibility criteria for the non-OUD group: no history of substance use disorders (excluding cannabis or nicotine); no being treated with medication for OUD. MRI was performed on a 3T Siemens Prisma scanner. For fMRI, T2*-weighted BOLD images were acquired using a single-shot gradient-echo echo-planar imaging sequence with the following parameters: repetition time (TR)=800 ms, echo time (TE)=37 ms, flip angle (FA)=52°, field of view (FOV)=208×208 mm², voxel size=2×2×2 mm³, 72 slices parallel to the anterior-posterior commissural line, scan duration=470.4 sec, multi-band acceleration factor=8. For structural MRI, images were acquired using the magnetization-prepared rapid gradient-echo imaging sequence and prospective motion correction with volumetric navigators with the following parameters: TR/TE=2430/2.24 ms, FA=8°, FOV=240×256 mm², voxel size=0.8×0.8×0.8 mm³, 208 sagittal slices.

**Study 2** (11 OUD, 8 non-OUD). Eligibility criteria for both groups: 18–50 years of age; no MRI contraindications; no comorbid central nervous system disease that might interfere with study procedures; no chronic use of anti-convulsant medications except gabapentin. Additional eligibility criteria for the OUD group: DSM-IV-TR diagnosis of opioid dependence or DSM-5 diagnosis of OUD; treatment with a stable dosage of medication for OUD for at least 2 years. Additional eligibility criteria for the non-OUD group: no diagnosis of opioid dependence or OUD. MRI was performed on a 3T Siemens Prisma scanner. For fMRI, T2*-weighted BOLD images were acquired using a single-shot gradient-echo echo-planar imaging sequence with the following parameters: TR/TE=800/37 ms, FA=52°, FOV=208×208 mm², voxel size=2×2×2 mm³, 72 slices parallel to the anterior-posterior commissural line, scan duration=350.4 sec, multi-band acceleration factor=8. For structural MRI, images were acquired using the magnetization-prepared rapid gradient-echo imaging sequence with the following parameters: TR/TE=2400/2.24 ms, FA=8°, FOV=240×256 mm², voxel size=0.8×0.8×0.8 mm³, 208 sagittal slices.

**Study 3** (9 OUD, 2 non-OUD). Eligibility criteria for both groups: 18–60 years of age; no claustrophobia or other MRI contraindications; working command of English; a negative HIV test at screening; not currently pregnant or breastfeeding; body weight ≤350 lbs; no history of epilepsy or seizure disorder; no current serious psychiatric disorder that compromise successful participation in the study; no self-reported heavy daily use of psychoactive substances. Additional eligibility criteria for the OUD group: DSM-5 diagnosis of OUD; treatment with a stable dosage of medication for OUD. Additional eligibility criteria for the non-OUD group: no history of substance use disorders (excluding cannabis or nicotine); no opioid use in the past 30 days; no positive urine drug tests for illicit substances. MRI was performed on a 3T Siemens Prisma scanner. For fMRI, T2*-weighted BOLD images were acquired using a single-shot gradient-echo echo-planar imaging sequence with the following parameters: TR/TE=800/37 ms, FA=52°, FOV=208×208 mm², voxel size=2×2×2 mm³, 72 slices parallel to the anterior-posterior commissural line, scan duration=470.4 sec, multi-band acceleration factor=8. For structural MRI, images were acquired using the magnetization-prepared rapid gradient-echo imaging sequence with the following parameters: TR/TE=2400/2.24 ms, FA=8°, FOV=240×256 mm², voxel size=0.8×0.8×0.8 mm³, 208 sagittal slices.

**Study 4** (74 OUD, 49 non-OUD). Eligibility criteria for both groups: 18–60 years of age; no claustrophobia or other MRI contraindications; good physical health ascertained by history and physical examination, blood chemistry and urinalysis; no current use of medications that may confound blood oxygen level-dependent brain response; no current psychosis, dementia, intellectual disability, or history of schizophrenia; no clinically significant cardiovascular, hematologic, pulmonary, hepatic, renal, metabolic, gastrointestinal, neurologic, or endocrine abnormalities; not currently pregnant or breastfeeding; no history of clinically significant head trauma. Additional eligibility criteria for the OUD group: DSM-IV-TR diagnosis of opioid dependence confirmed by self-report and medical records documenting daily opioid use for more than 2 weeks in the past 3 months; evidence of detoxification from opioids as established by urine drug screens and a negative naloxone challenge test; no contraindications for extended-release naltrexone. Additional eligibility criteria for the non-OUD group: no diagnosis of opioid dependence; current tobacco use. MRI was performed on a 3T Siemens Tim Trio scanner. For fMRI, T2*-weighted BOLD images were acquired using a single-shot gradient-echo echo-planar imaging sequence with the following parameters: TR=2000/3000 ms, TE=30/32 ms, FA=90°, FOV=220×220/192×192 mm², voxel size=3.4×3.4×4.5/3×3×3 mm³, 32/46 slices parallel to the anterior-posterior commissural line, scan duration=290/360 sec. For structural MRI, images were acquired using the magnetization-prepared rapid gradient-echo imaging sequence with the following parameters: TR/TE=1510/3.71 ms, FA=9°, FOV=192×256 mm², voxel size=1×1×1 mm³, 160 sagittal slices.

**Study 5** (34 OUD, 16 non-OUD). Eligibility criteria for both groups: 18–42 years of age; pre-menopausal females who have given birth to at least one child 3 years old or younger; no claustrophobia or other MRI contraindications; not currently pregnant; no history of significant head trauma or brain surgery; no clinically significant medical disorder or condition that may affect cerebral function or circulation; no current DSM-IV-TR Axis 1 disorder such as schizophrenia, major depression, or psychosis; no current DSM-IV-TR Axis 2 disorder such as cognitive impairment or severe psychopathology that may compromise successful participation in the study. Additional eligibility criteria for the OUD group: DSM-IV-TR diagnosis of opioid dependence; treatment with a stable dosage of medication for OUD. Additional eligibility criteria for the non-OUD group: no history of drug dependence. MRI was performed on a 3T Siemens Prisma scanner. For fMRI, T2*-weighted BOLD images were acquired using a single-shot gradient-echo echo-planar imaging sequence with the following parameters: TR/TE=500/25 ms, FA=30°, FOV=192×192 $mm^2$, voxel size=3×3×3 $mm^3$, 48 slices parallel to the anterior-posterior commissural line, scan duration=590 sec, multi-band acceleration factor=6. For structural MRI, images were acquired using the magnetization-prepared rapid gradient-echo imaging sequence with the following parameters: TR/TE=182/3.51 ms, FA=9°, FOV=220×160 $mm^2$, voxel size=0.9×0.9×1.0 $mm^3$, 160 sagittal slices.

**Study 6** (8 OUD, 15 non-OUD). Eligibility criteria for both groups: 18-50 years of age; no claustrophobia or other MRI contraindications; no psychotropic medication that interferes with opioid receptor binding; no severe substance use disorder other than OUD or cannabis use disorder; no allergy to any opioid or naloxone; no history of seizure disorder, head trauma, or brain tumor; not currently pregnant or breastfeeding; breath alcohol concentration lower than 0.01%; no positive urine drug tests for illicit substances; no medical condition, illness, or disorder that may compromise participant safety or successful participation in the study. Additional eligibility criteria for the OUD group: DSM-5 diagnosis of OUD; treatment with a stable dosage of medication for OUD. Additional eligibility criteria for the non-OUD group: no history of OUD; no opioid use in the past 30 days; no major psychiatric diagnosis; no substance use disorder or treatment of a substance use disorder; alcohol consumption less than 15 standard drinks per week for men and less than 8 for women. MRI was performed on a 3T Siemens Prisma scanner. For fMRI, T2*-weighted BOLD images were acquired using a single-shot gradient-echo echo-planar imaging sequence and prospective motion correction with volumetric navigators with the following parameters: TR/TE=4180/31 ms, FA=90°, FOV=220×220 $mm^2$, voxel size=2×2×2 $mm^3$, 61 slices parallel to the anterior-posterior commissural line, scan duration=614.5 sec. For structural MRI, images were acquired using the multi-echo magnetization-prepared rapid gradient-echo imaging sequence with the following parameters: TR=2500 ms, TE=1.86, 3.78, 5.70, 7.62 ms, FA=8°, FOV=240×256 $mm^2$, voxel size=0.8×0.8×0.8 $mm^3$, 224 sagittal slices.

**Study 7** (2 OUD, 12 non-OUD). Eligibility criteria for both groups: 18–65 years of age; no claustrophobia or other MRI contraindications; not currently pregnant or breastfeeding; body weight ≤350 lbs; no history of epilepsy or seizure disorder; history of head trauma; no organ dysfunction; no history of schizophrenia or psychotic disorder; no current bipolar disorder, eating disorder, or major depression with suicidal ideation or psychotic features; alcohol consumption less than 15 standard drinks per week; no medical condition, illness, or disorder that may compromise participant safety or successful participation in the study. Additional eligibility criteria for the OUD group: DSM-5 diagnosis of OUD; treatment with a stable dosage of medication for OUD for at least four weeks. Additional eligibility criteria for the non-OUD group: no history of OUD; no opioid use in the past 30 days. MRI was performed on a 3T Siemens Prisma scanner. For fMRI, T2*-weighted BOLD images were acquired using a single-shot gradient-echo echo-planar imaging sequence with the following parameters: TR/TE=4180/31 ms, FA=90°, FOV=220×220 $mm^2$, voxel size=2×2×2 $mm^3$, 61 slices parallel to the anterior-posterior commissural line, scan duration=614.5 sec. For structural MRI, images were acquired using the multi-echo magnetization-prepared rapid gradient-echo imaging sequence and prospective motion correction with volumetric navigators with the following parameters: TR=2500 ms, TE=1.86, 3.78, 5.70, 7.62 ms, FA=8°, FOV=240×256 $mm^2$, voxel size=0.8×0.8×0.8 $mm^3$, 224 sagittal slices.

***Exploratory graph-theoretical analyses***

Exploratory graph-theoretical analyses were conducted in R. For each participant, the adjacency matrix was binarized across a range of sparsity thresholds (0.09–0.50, in 0.01 increments) to minimize the confounding effect of individual differences in mean global connectivity strength and reduce potential bias associated with selecting a single threshold.[1] Thresholds were selected such that the largest connected component contained at least 50% of all brain regions (i.e., nodes), while no more than 50% of all possible interregional connections (i.e., edges) were retained. At each threshold, brain network partitions were estimated using the Louvain algorithm.[2] Due to the stochastic nature of Louvain optimization, the algorithm was repeated 100 times for each thresholded matrix. Modularity ($Q$) was computed for each of the 100 partitions, and the highest $Q$ was retained.[3] A higher $Q$ value indicates a greater extent to which a network can be subdivided into internally dense and externally sparse communities, i.e., stronger segregation. To quantify the similarity between the canonical Power-264 atlas partition and the empirically derived partition, normalized mutual information ($NMI$) was calculated using the partition associated with the highest $Q$.[4] A higher $NMI$ value indicates greater correspondence between the empirical and canonical network organization. Global efficiency $E_g$was also computed from the same partition to assess whole-brain integration, with higher $E_g$ values indicating greater network integration.[5] The computations of $Q$, $NMI$, and $E_g$ are as follows:

$$Q = \frac{1}{2M} \sum\nolimits_{i \neq j} (A_{i,j} - \frac{k_i k_j}{2M}) \delta_{i,j} \ ,$$

$$NMI = \frac{2 \sum_p \sum_q n_{p,q} \log_2 (\frac{n_{p,q} N}{n_p n_q})}{- \sum_p n_p \log_2 \left( \frac{n_p}{N} \right) - \sum_q q \log_2 \left( \frac{n_q}{N} \right)} ,$$

$$E_g = \frac{1}{N(N-1)} \sum\nolimits_{i \neq j} \frac{1}{d_{i,j}} ,$$

where $M$ is the total number of edges; $A_{i,j}$ is the binary connection between nodes $i$ and $j$; $k_i$ and $k_j$ are the degree of nodes $i$ and $j$, respectively; $\delta_{i,j}$ is an indicator denoting whether nodes $i$ and $j$ belong to the same community; $N$ is the total number of nodes; $n_p$ is the number of nodes in the $p$-th network of the Power-264 atlas; $n_q$ is the number of nodes in the $q$-th network of the empirically derived partition; $n_{p,q}$ is the number of nodes shared by both the $p$-th Power-24 network and the $q$-th empirical network; and $d_{i,j}$ is the shortest path length between nodes $i$ and $j$. For each participant, $Q$, $NMI$, and $E_g$ values were averaged across sparsity thresholds to derive a single summary estimate for statistical analyses.

### SUPPLEMENTARY RESULTS

#### *Effects of sex, overdose, and medication*

The effect of sex and the interaction between sex and group were not significant on the segregation of the association network or the sensorimotor network (p=0.18–0.79) (see **Table S5** and **Table S6**).

There was a significant effect of past opioid overdose on brain network segregation for the association network ($F(1,47.00)=5.89$, $p=0.019$), such that patients with a history of overdose had higher segregation (0.29 [0.23,0.35]) than those without such a history (0.24 [0.18,0.30]). The effect of overdose on the segregation of the sensorimotor network was not significant (p=0.17) (see **Table S7**).

In terms of medication for OUD, patients treated with methadone and those treated with buprenorphine did not significantly differ in brain network segregation of either the association network or the sensorimotor network (p=0.11 & 0.31) (see **Table S8**).

### SUPPLEMENTARY TABLES

***Table S1. Main effect of group.***

| Variable | F | $df_1, df_2$ | p | Partial $\eta^2$ | OUD | Non-OUD |
|---|---|---|---|---|---|---|
| ***Segregation of the association network*** | | | | | | |
| Overall | 12.27 | 1, 241.57 | <0.001 | 0.05 | 0.25 [0.22,0.27] | 0.29 [0.26,0.31] |
| Cingulo-opercular | 9.30 | 1, 232.51 | 0.003 | 0.04 | 0.25 [0.21,0.29] | 0.30 [0.26,0.35] |
| Dorsal attention | 0.01 | 1, 245.53 | 0.92 | 0.00 | 0.30 [0.26,0.35] | 0.30 [0.25,0.34] |
| Default mode | 6.27 | 1, 255.00 | 0.013 | 0.02 | 0.30 [0.26,0.33] | 0.34 [0.30,0.38] |
| Fronto-parietal | 2.35 | 1, 235.60 | 0.13 | 0.01 | 0.26 [0.22,0.30] | 0.28 [0.24,0.32] |
| Salience | 3.00 | 1, 243.33 | 0.084 | 0.01 | 0.24 [0.20,0.28] | 0.27 [0.23,0.32] |
| Ventral attention | 12.42 | 1, 255.00 | <0.001 | 0.05 | 0.14 [0.09,0.19] | 0.22 [0.17,0.28] |
| ***Segregation of the sensorimotor network*** | | | | | | |
| Overall | 13.14 | 1, 209.83 | <0.001 | 0.06 | 0.41 [0.37,0.45] | 0.46 [0.42,0.50] |
| Auditory | 2.70 | 1, 255.00 | 0.10 | 0.01 | 0.30 [0.26,0.34] | 0.33 [0.29,0.37] |
| Hand | 10.76 | 1, 242.41 | 0.001 | 0.04 | 0.32 [0.28,0.36] | 0.38 [0.33,0.42] |
| Mouth | 2.48 | 1, 173.05 | 0.12 | 0.01 | 0.52 [0.36,0.68] | 0.57 [0.41,0.73] |
| Visual | 5.93 | 1, 239.85 | 0.016 | 0.02 | 0.50 [0.46,0.54] | 0.54 [0.50,0.59] |
| ***Segregation of other networks*** | | | | | | |
| Cerebellar | 2.22 | 1, 243.33 | 0.14 | 0.01 | 0.18 [−0.02,0.39] | 0.31 [0.10,0.51] |
| Memory | 0.50 | 1, 237.78 | 0.48 | 0.00 | 0.41 [0.32,0.50] | 0.39 [0.29,0.48] |
| Subcortical | 10.01 | 1, 209.12 | 0.002 | 0.05 | 0.13 [0.04,0.23] | 0.21 [0.11,0.31] |

**Abbreviation:** df, degree of freedom; OUD, opioid use disorder.

***Table S2. Main effect of years of opioid use.***

| Variable | F | $df_1, df_2$ | p | Partial $\eta^2$ | Slope ($\times 10^{-3}$) |
|---|---|---|---|---|---|
| ***Segregation of the association network*** | | | | | |
| Overall | 4.05 | 1, 115.00 | 0.046 | 0.03 | −2.21 [−4.27,−0.15] |
| Cingulo-opercular | 0.01 | 1, 114.04 | 0.91 | 0.00 | −0.21 [−3.64,3.72] |
| Dorsal attention | 4.56 | 1, 114.20 | 0.035[a] | 0.04 | −3.93 [−7.58,−0.70] |
| Default mode | 4.87 | 1, 115.00 | 0.029[a] | 0.04 | −3.69 [−6.84,−0.55] |
| Fronto-parietal | 3.38 | 1, 114.11 | 0.069 | 0.03 | −2.83 [−6.06,−0.28] |
| Salience | 0.18 | 1, 115.00 | 0.67 | 0.00 | 0.78 [−2.70,4.25] |
| Ventral attention | 1.67 | 1, 115.00 | 0.20 | 0.01 | −3.05 [−7.48,1.38] |
| ***Segregation of the sensorimotor network*** | | | | | |
| Overall | 1.25 | 1, 114.25 | 0.27 | 0.01 | 1.58 [−0.66,4.72] |
| Auditory | 1.92 | 1, 112.83 | 0.17 | 0.02 | 2.81 [−0.69,6.92] |
| Hand | 0.67 | 1, 113.77 | 0.41 | 0.01 | 1.53 [−1.73,5.25] |
| Mouth | 0.78 | 1, 113.50 | 0.38 | 0.01 | 2.88 [−3.54,8.78] |
| Visual | 0.01 | 1, 115.00 | 0.94 | 0.00 | 0.14 [−3.41,3.68] |
| ***Segregation of other networks*** | | | | | |
| Cerebellar | 0.00 | 1, 115.00 | >0.99 | 0.00 | 0.02 [−8.51,8.56] |
| Memory | 0.12 | 1, 115.00 | 0.73 | 0.00 | 1.07 [−4.78,6.93] |
| Subcortical | 0.51 | 1, 114.50 | 0.48 | 0.00 | 1.57 [−3.12,5.43] |

**Abbreviation:** df, degree of freedom.
[a] Significance did not survive correction false discovery rate (FDR).

***Table S3. Main effect of age (years).***

| Variable | F | $df_1, df_2$ | p | Partial $\eta^2$ | Slope ($\times10^{-3}$) |
|---|---|---|---|---|---|
| ***Segregation of the association network*** | | | | | |
| Overall | 20.19 | 1, 134.41 | <0.001 | 0.13 | −2.14 [−3.09,−1.28] |
| Cingulo-opercular | 23.73 | 1, 97.48 | <0.001 | 0.20 | −3.70 [−5.17,−2.29] |
| Dorsal attention | 11.54 | 1, 165.14 | <0.001 | 0.07 | −2.69 [−4.29,−1.30] |
| Default mode | 6.46 | 1, 255.00 | 0.012 | 0.02 | −1.85 [−3.25,−0.45] |
| Fronto-parietal | 2.52 | 1, 112.49 | 0.12 | 0.02 | −1.12 [−2.45,0.22] |
| Salience | 2.73 | 1, 174.60 | 0.10 | 0.02 | −1.27 [−2.82,0.05] |
| Ventral attention | 5.19 | 1, 255.00 | 0.024 | 0.02 | −2.23 [−4.12,−0.35] |
| ***Segregation of the sensorimotor network*** | | | | | |
| Overall | 9.89 | 1, 201.85 | 0.002 | 0.05 | −1.90 [−2.62,−0.37] |
| Auditory | 3.79 | 1, 255.00 | 0.053 | 0.01 | −1.55 [−3.08,−0.02] |
| Hand | 1.69 | 1, 175.84 | 0.20 | 0.01 | −1.00 [−2.28,0.60] |
| Mouth | 8.63 | 1, 254.15 | 0.004 | 0.03 | −3.80 [−5.38,0.12] |
| Visual | 2.82 | 1, 121.50 | 0.096 | 0.02 | −1.31 [−2.74,0.21] |
| ***Segregation of other networks*** | | | | | |
| Cerebellar | 2.55 | 1, 184.39 | 0.11 | 0.01 | 5.64 [−2.02,11.16] |
| Memory | 0.33 | 1, 238.23 | 0.57 | 0.00 | 0.77 [−2.07,3.14] |
| Subcortical | 8.16 | 1, 254.66 | 0.005 | 0.03 | −2.85 [−4.88,−1.02] |

**Abbreviation:** df, degree of freedom.

***Table S4. Interaction between group and age (years).***

| Variable | F | $df_1, df_2$ | p | Partial $\eta^2$ | OUD slope ($\times10^{-3}$) | Non-OUD slope ($\times10^{-3}$) |
|---|---|---|---|---|---|---|
| ***Segregation of the association network*** | | | | | | |
| Overall | 0.13 | 1, 154.26 | 0.72 | 0.00 | −2.30 [−3.74,−0.86] | −1.97 [−3.33,−0.62] |
| Cingulo-opercular | 0.26 | 1, 120.40 | 0.61 | 0.00 | −4.06 [−6.35,−1.76] | −3.31 [−5.47,−1.15] |
| Dorsal attention | 0.45 | 1, 175.56 | 0.50 | 0.00 | −3.21 [−5.59,−0.83] | −2.18 [−4.43,0.06] |
| Default mode | 0.01 | 1, 254.00 | 0.91 | 0.00 | −1.93 [−4.20,0.34] | −1.77 [−3.88,0.34] |
| Fronto-parietal | 0.37 | 1, 126.44 | 0.54 | 0.00 | −0.72 [−2.85,1.41] | −1.55 [−3.55,0.45] |
| Salience | 0.13 | 1, 191.57 | 0.72 | 0.00 | −1.53 [−3.80,0.74] | −1.00 [−3.15,1.16] |
| Ventral attention | 0.04 | 1, 254.00 | 0.84 | 0.00 | −2.42 [−5.47,0.63] | −2.02 [−4.86,0.81] |
| ***Segregation of the sensorimotor network*** | | | | | | |
| Overall | 1.17 | 1, 241.65 | 0.28 | 0.00 | −2.65 [−4.38,−0.92] | −1.39 [−3.03,0.25] |
| Auditory | 3.24 | 1, 157.69 | 0.074 | 0.02 | −2.93 [−5.36,−0.49] | −0.13 [−2.41,2.15] |
| Hand | 0.35 | 1, 199.64 | 0.55 | 0.00 | −1.45 [−3.72,0.82] | −0.56 [−2.72,1.59] |
| Mouth | 0.33 | 1, 252.49 | 0.57 | 0.00 | −4.57 [−8.23,−0.91] | −3.15 [−6.60,0.30] |
| Visual | 0.25 | 1, 144.12 | 0.62 | 0.00 | −0.94 [−3.30,1.42] | −1.69 [−3.90,0.53] |
| ***Segregation of other networks*** | | | | | | |
| Cerebellar | 1.20 | 1, 212.90 | 0.27 | 0.01 | 9.46 [−0.89,19.82] | 1.98 [−7.86,11.81] |
| Memory | 1.74 | 1, 233.32 | 0.19 | 0.01 | −1.01 [−4.88,2.85] | 2.39 [−1.28,6.06] |
| Subcortical | 0.33 | 1, 253.67 | 0.57 | 0.00 | −3.43 [−6.27,−0.59] | −2.33 [−5.01,0.35] |

**Abbreviation:** df, degree of freedom; OUD, opioid use disorder.

***Table S5. Main effect of sex.***

| Variable | F | $df_1, df_2$ | p | Partial $\eta^2$ | Male | Female |
|---|---|---|---|---|---|---|
| *Segregation of the association network* | | | | | | |
| Overall | 1.79 | 1, 132.80 | 0.18 | 0.01 | 0.26 [0.23,0.29] | 0.27 [0.25,0.30] |
| Cingulo-opercular | 1.22 | 1, 95.92 | 0.27 | 0.01 | 0.27 [0.22,0.31] | 0.28 [0.25,0.32] |
| Dorsal attention | 0.01 | 1, 163.74 | 0.93 | 0.00 | 0.30 [0.25,0.35] | 0.30 [0.26,0.34] |
| Default mode | 3.78 | 1, 255.00 | 0.053 | 0.01 | 0.30 [0.26,0.34] | 0.33 [0.29,0.37] |
| Fronto-parietal | 2.99 | 1, 110.95 | 0.087 | 0.03 | 0.26 [0.22,0.30] | 0.28 [0.25,0.32] |
| Salience | 0.05 | 1, 173.11 | 0.83 | 0.00 | 0.26 [0.21,0.30] | 0.25 [0.21,0.30] |
| Ventral attention | 0.33 | 1, 255.00 | 0.56 | 0.00 | 0.18 [0.12,0.23] | 0.19 [0.14,0.23] |
| *Segregation of the sensorimotor network* | | | | | | |
| Overall | 0.07 | 1, 195.12 | 0.79 | 0.00 | 0.44 [0.40,0.48] | 0.44 [0.40,0.47] |
| Auditory | 3.33 | 1, 255.00 | 0.069 | 0.01 | 0.33 [0.28,0.37] | 0.30 [0.26,0.34] |
| Hand | 0.65 | 1, 174.24 | 0.42 | 0.00 | 0.36 [0.31,0.40] | 0.34 [0.30,0.39] |
| Mouth | 1.38 | 1, 254.72 | 0.24 | 0.01 | 0.53 [0.37,0.69] | 0.56 [0.40,0.72] |
| Visual | 0.02 | 1, 119.78 | 0.88 | 0.00 | 0.52 [0.48,0.57] | 0.52 [0.48,0.56] |
| *Segregation of other networks* | | | | | | |
| Cerebellar | 1.58 | 1, 182.78 | 0.21 | 0.01 | 0.29 [0.08,0.50] | 0.20 [0.01,0.40] |
| Memory | 1.05 | 1, 235.21 | 0.31 | 0.00 | 0.38 [0.29,0.48] | 0.41 [0.32,0.50] |
| Subcortical | 0.09 | 1, 252.76 | 0.77 | 0.00 | 0.17 [0.07,0.27] | 0.17 [0.07,0.27] |

**Abbreviation:** df, degree of freedom.

***Table S6. Interaction between group and sex.***

| Variable | F | $df_1, df_2$ | p | Partial $\eta^2$ | OUD male | OUD female | Non-OUD male | Non-OUD female |
|---|---|---|---|---|---|---|---|---|
| *Segregation of the association network* | | | | | | | | |
| Overall | 0.51 | 1, 254.96 | 0.48 | 0.00 | 0.23 [0.20,0.27] | 0.25 [0.21,0.28] | 0.28 [0.25,0.32] | 0.29 [0.25,0.32] |
| Cingulo-opercular | 0.06 | 1, 254.97 | 0.81 | 0.00 | 0.23 [0.17,0.28] | 0.24 [0.19,0.30] | 0.29 [0.24,0.35] | 0.30 [0.25,0.36] |
| Dorsal attention | 0.12 | 1, 255.00 | 0.72 | 0.00 | 0.29 [0.24,0.35] | 0.29 [0.24,0.34] | 0.30 [0.24,0.35] | 0.30 [0.25,0.35] |
| Default mode | 0.29 | 1, 255.00 | 0.59 | 0.00 | 0.28 [0.23,0.32] | 0.30 [0.26,0.34] | 0.32 [0.27,0.37] | 0.36 [0.31,0.40] |
| Fronto-parietal | 0.09 | 1, 254.84 | 0.76 | 0.00 | 0.24 [0.20,0.29] | 0.27 [0.23,0.31] | 0.27 [0.23,0.32] | 0.29 [0.25,0.33] |
| Salience | 1.66 | 1, 255.00 | 0.20 | 0.01 | 0.23 [0.18,0.28] | 0.24 [0.20,0.29] | 0.28 [0.23,0.34] | 0.26 [0.22,0.31] |
| Ventral attention | 2.33 | 1, 254.87 | 0.13 | 0.01 | 0.12 [0.06,0.18] | 0.15 [0.09,0.21] | 0.24 [0.17,0.30] | 0.22 [0.16,0.27] |
| *Segregation of the sensorimotor network* | | | | | | | | |
| Overall | 0.77 | 1, 255.00 | 0.38 | 0.00 | 0.41 [0.37,0.45] | 0.40 [0.36,0.43] | 0.46 [0.42,0.50] | 0.46 [0.43,0.49] |
| Auditory | 0.50 | 1, 255.00 | 0.48 | 0.00 | 0.31 [0.26,0.36] | 0.28 [0.23,0.32] | 0.34 [0.28,0.39] | 0.32 [0.28,0.37] |
| Hand | 0.43 | 1, 254.88 | 0.51 | 0.00 | 0.32 [0.27,0.37] | 0.32 [0.27,0.36] | 0.39 [0.34,0.44] | 0.37 [0.32,0.41] |
| Mouth | 1.36 | 1, 255.00 | 0.24 | 0.01 | 0.51 [0.43,0.59] | 0.51 [0.43,0.59] | 0.56 [0.48,0.65] | 0.62 [0.54,0.69] |
| Visual | 0.42 | 1, 255.00 | 0.52 | 0.00 | 0.50 [0.45,0.55] | 0.49 [0.44,0.53] | 0.54 [0.49,0.59] | 0.55 [0.50,0.59] |
| *Segregation of other networks* | | | | | | | | |
| Cerebellar | 0.34 | 1, 254.83 | 0.56 | 0.00 | 0.26 [0.05,0.48] | 0.15 [−0.06,0.36] | 0.34 [0.11,0.56] | 0.29 [0.09,0.49] |
| Memory | 0.23 | 1, 254.90 | 0.63 | 0.00 | 0.39 [0.30,0.49] | 0.43 [0.34,0.52] | 0.38 [0.28,0.48] | 0.40 [0.30,0.49] |
| Subcortical | 0.70 | 1, 253.98 | 0.40 | 0.00 | 0.12 [0.01,0.22] | 0.13 [0.03,0.24] | 0.21 [0.11,0.32] | 0.20 [0.10,0.31] |

**Abbreviation:** df, degree of freedom; OUD, opioid use disorder.

***Table S7. Main effect of opioid overdose history.***

| Variable | F | $df_1, df_2$ | p | Partial $\eta^2$ | History of overdose | Never overdose |
|---|---|---|---|---|---|---|
| ***Segregation of the association network*** | | | | | | |
| Overall | 5.89 | 1, 47.00 | 0.019 | 0.11 | 0.29 [0.23,0.35] | 0.24 [0.18,0.30] |
| Cingulo-opercular | 1.02 | 1, 47.00 | 0.32 | 0.02 | 0.24 [0.11,0.36] | 0.19 [0.07,0.32] |
| Dorsal attention | 0.80 | 1, 46.80 | 0.37 | 0.02 | 0.32 [0.20,0.44] | 0.29 [0.17,0.41] |
| Default mode | 2.93 | 1, 47.00 | 0.094 | 0.06 | 0.40 [0.26,0.54] | 0.34 [0.21,0.48] |
| Fronto-parietal | 1.38 | 1, 45.96 | 0.25 | 0.03 | 0.26 [0.07,0.45] | 0.23 [0.04,0.42] |
| Salience | 4.01 | 1, 47.00 | 0.051 | 0.08 | 0.30 [0.19,0.41] | 0.22 [0.11,0.33] |
| Ventral attention | 0.93 | 1, 47.00 | 0.34 | 0.02 | 0.19 [0.06,0.32] | 0.14 [0.01,0.27] |
| ***Segregation of the sensorimotor network*** | | | | | | |
| Overall | 1.94 | 1, 45.65 | 0.17 | 0.04 | 0.41 [0.33,0.50] | 0.37 [0.29,0.46] |
| Auditory | 0.47 | 1, 47.00 | 0.50 | 0.01 | 0.22 [0.11,0.34] | 0.25 [0.13,0.37] |
| Hand | 0.00 | 1, 46.81 | 0.96 | 0.00 | 0.28 [0.14,0.42] | 0.28 [0.14,0.41] |
| Mouth | 4.09 | 1, 44.77 | 0.049[a] | 0.08 | 0.60 [0.40,0.80] | 0.46 [0.26,0.67] |
| Visual | 1.71 | 1, 47.00 | 0.20 | 0.04 | 0.54 [0.44,0.64] | 0.50 [0.39,0.60] |
| ***Segregation of other networks*** | | | | | | |
| Cerebellar | 0.13 | 1, 47.00 | 0.72 | 0.00 | 0.15 [−0.15,0.44] | 0.18 [−0.11,0.48] |
| Memory | 2.02 | 1, 47.00 | 0.16 | 0.04 | 0.48 [0.29,0.67] | 0.39 [0.20,0.58] |
| Subcortical | 2.16 | 1, 46.99 | 0.15 | 0.04 | 0.21 [−0.02,0.44] | 0.14 [−0.09,0.36] |

**Abbreviation:** df, degree of freedom.
[a] Significance did not survive correction false discovery rate (FDR).

***Table S8. Main effect of medication.***

| Variable | F | $df_1, df_2$ | p | Partial $\eta^2$ | Methadone | Buprenorphine |
|---|---|---|---|---|---|---|
| ***Segregation of the association network*** | | | | | | |
| Overall | 2.58 | 1, 60.00 | 0.11 | 0.04 | 0.25 [0.19,0.31] | 0.28 [0.23,0.34] |
| Cingulo-opercular | 0.20 | 1, 60.00 | 0.66 | 0.00 | 0.25 [0.16,0.34] | 0.26 [0.18,0.34] |
| Dorsal attention | 1.60 | 1, 58.57 | 0.21 | 0.03 | 0.31 [0.18,0.43] | 0.34 [0.22,0.47] |
| Default mode | 1.02 | 1, 60.00 | 0.32 | 0.02 | 0.28 [0.18,0.38] | 0.31 [0.22,0.40] |
| Fronto-parietal | 1.07 | 1, 60.00 | 0.31 | 0.02 | 0.27 [0.20,0.34] | 0.29 [0.23,0.36] |
| Salience | 0.16 | 1, 57.94 | 0.69 | 0.00 | 0.26 [0.16,0.37] | 0.27 [0.18,0.37] |
| Ventral attention | 2.72 | 1, 60.00 | 0.10 | 0.04 | 0.15 [0.03,0.27] | 0.21 [0.10,0.33] |
| ***Segregation of the sensorimotor network*** | | | | | | |
| Overall | 1.07 | 1, 60.00 | 0.31 | 0.02 | 0.45 [0.38,0.51] | 0.47 [0.41,0.53] |
| Auditory | 0.87 | 1, 60.00 | 0.35 | 0.01 | 0.31 [0.20,0.41] | 0.34 [0.24,0.43] |
| Hand | 0.06 | 1, 59.13 | 0.81 | 0.00 | 0.38 [0.28,0.47] | 0.37 [0.28,0.46] |
| Mouth | 0.04 | 1, 58.83 | 0.85 | 0.00 | 0.60 [0.48,0.71] | 0.59 [0.48,0.70] |
| Visual | 5.80 | 1, 60.00 | 0.019[a] | 0.09 | 0.51 [0.41,0.60] | 0.58 [0.49,0.67] |
| ***Segregation of other network*** | | | | | | |
| Cerebellar | 0.15 | 1, 60.00 | 0.70 | 0.00 | 0.25 [−0.48,0.97] | 0.16 [−0.51,0.82] |
| Memory | 0.75 | 1, 57.14 | 0.39 | 0.01 | 0.50 [0.22,0.78] | 0.45 [0.17,0.72] |
| Subcortical | 2.47 | 1, 58.96 | 0.12 | 0.04 | 0.18 [0.03,0.33] | 0.12 [−0.02,0.26] |

**Abbreviation:** df, degree of freedom.
[a] Significance did not survive correction false discovery rate (FDR).

### SUPPLEMENTARY FIGURES

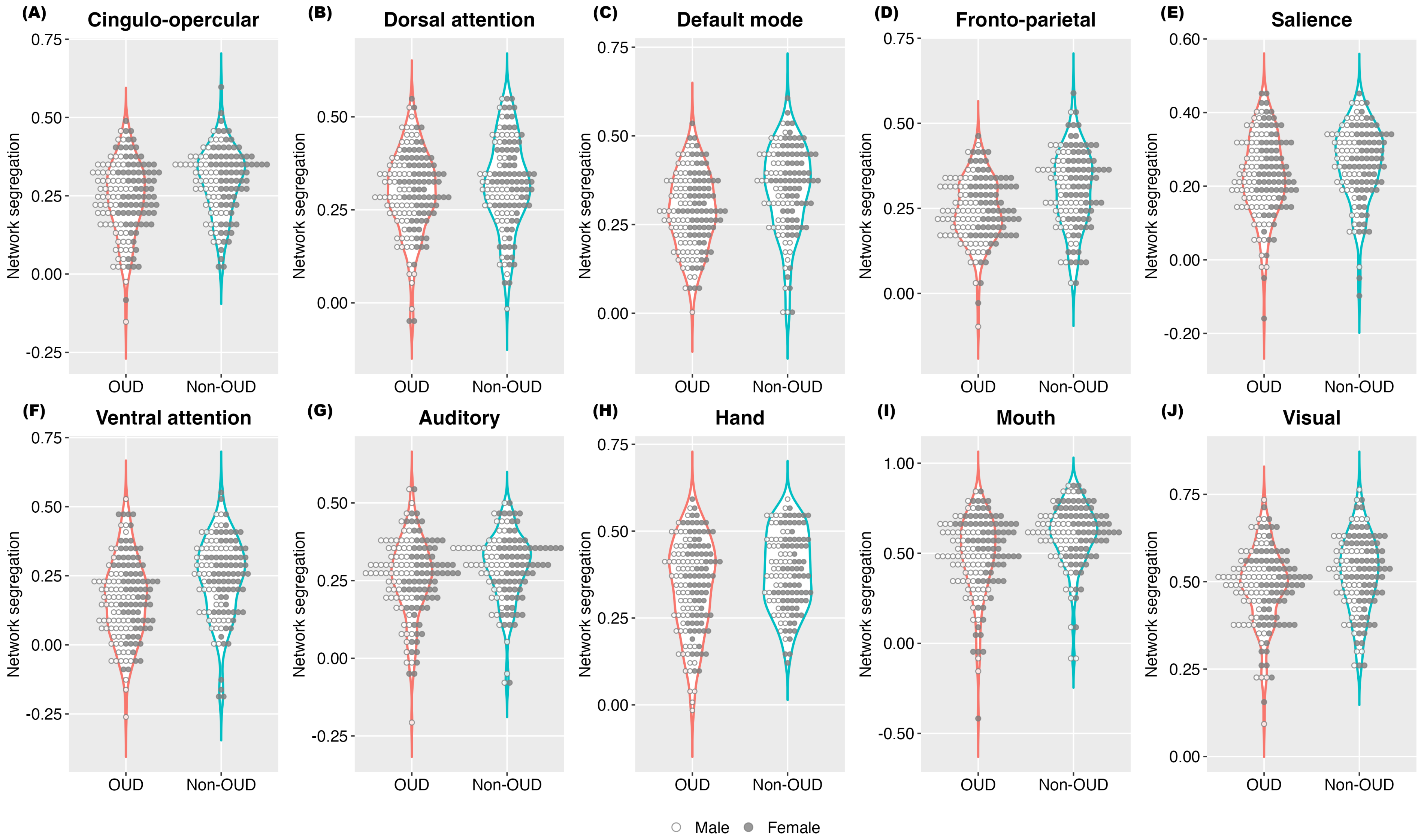


***Figure S1. Segregation of individual brain networks.***

Abbreviation: OUD, opioid use disorder.

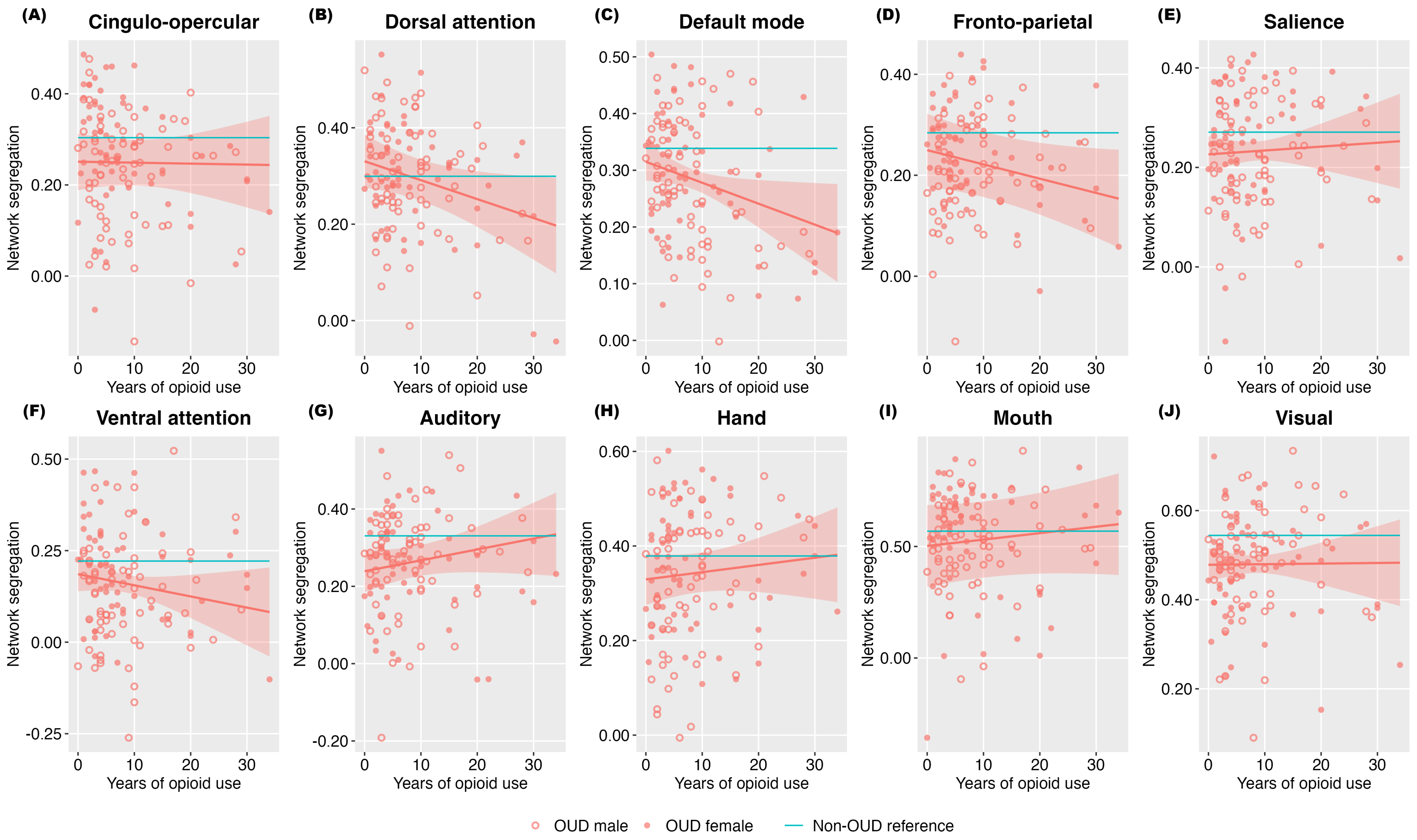


***Figure S2. Association between years of opioid use and segregation of individual brain networks.***

Shaded areas represent 95% confidence intervals. Abbreviation: OUD, opioid use disorder.

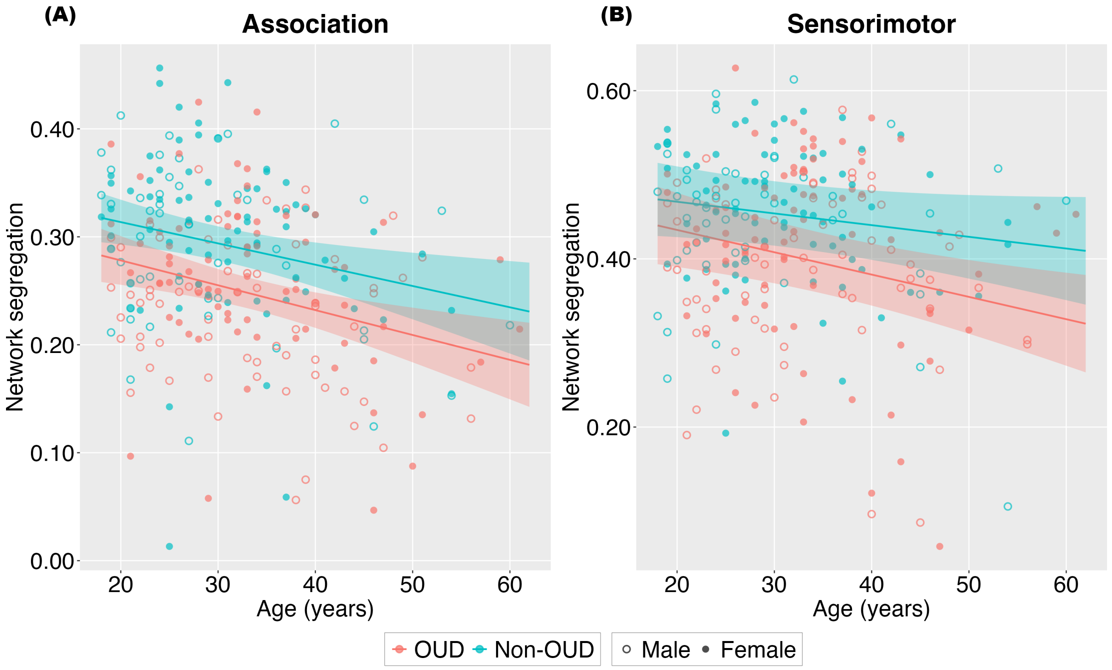


***Figure S3. Association between age and segregation of the association and sensorimotor networks.***

Shaded areas represent 95% confidence intervals. Abbreviation: OUD, opioid use disorder.
